# Assessing intention of linking to HIV services among female youth with diverse risk-taking profiles after interacting with an AI-powered tool in Gauteng

**DOI:** 10.64898/2026.11.27.26369061

**Authors:** Sinethemba Zamasishi Madlala, Simamkele Bokolo, Laura Rossouw, Caroline Govathson, Lawrence Long, Sarah Morris, Shawna Cooper, Sara Jewett, Candice Chetty-Makkan

## Abstract

**Introduction:** Adolescent girls and young women (AGYW) in South Africa face high HIV risk, yet engagement with HIV prevention and treatment services remains low. Digital health tools offer promising opportunities to support linkage to care, but their effectiveness may vary according to individual behavioural characteristics such as risk-taking propensity. Understanding how behavioural traits shape responses to AI-enabled tools is important for designing more targeted digital interventions. We assessed baseline risk-taking propensity among AGYW interacting with the Self Care from Anywhere (SCFA) platform and changes in intentions to engage in HIV services before and after interaction with the tool.

**Methods:** An observational pre-post design was conducted among 97 AGYW aged 16-24 years in Gauteng province (November 2024 to April 2025). Following informed consent, participants’ intention to link to HIV services was assessed before and after exposure to SCFA. Risk-taking propensity was measured using the Balloon Analogue Risk Task (BART), a validated behavioural measure that simulates real-world risk-taking, with higher scores indicating greater risk-taking propensity. Paired t-tests were used to examine pre-post changes in intention, while multivariate linear regression models estimated moderation effects while adjusting for socio-demographic and behavioural variables.

**Results:** Baseline BART scores varied among participants (M = 18.1; SD = 9.3), indicating diverse levels of risk-taking. After exposure to SCFA, intention to engage showed a slight increase, but the difference was not statistically significant (t (96) = 1.64, p = <0.105). There was a significant statistical interaction between post-intervention intention to engage in HIV services and baseline risk-taking (β = −0.313, p < 0.05), indicating that participants with lower risk-taking propensity (risk-averse) demonstrated an improvement in their intention to engage in HIV services. Age also moderated outcomes, with older participants’ intention to engage in HIV services increased more than younger participants (β = 0.372, p <0.05).

**Conclusion:** Interaction with SCFA increased intention to engage in HIV services, particularly among risk-averse and older AGYW. These findings suggest that AI-powered digital tools may influence users differently, based on behavioural characteristics, by allowing content, follow-ups prompts or referrals to be adapted to different risk profiles. This can be used to support HIV prevention strategies in a resource-limited setting.

## Introduction

Adolescent girls and young women (AGYW; aged 15-24 years) remain among the most affected populations by the HIV epidemic in sub-Saharan Africa [1,2]. Globally, women and girls of all ages accounted for approximately 45% of new infections in 2024, with the burden disproportionately concentrated in Sub-Saharan Africa (SSA), where 63% of new infections occurred among women and girls [1,3]. In contrast, in all other regions, approximately 73% of new infections occurred among men and boys, highlighting stark regional and gender disparities [1,3]. Among young people, the burden is particularly severe; an estimated 4 000 AGYW aged 15-24 years were newly infected with HIV each week in 2024, of whom approximately 3 300 were in sub-Saharan Africa [1,3]. In South Africa, AGYW account for nearly one-third of new infections annually [4]. Despite the expansion of HIV prevention strategies, many AGYW remain underserved due to structural inequalities, social stigma, and behavioural barriers that limit access to sexual and reproductive health services [4,5]. Although biomedical tools such as pre-exposure prophylaxis (PrEP) and HIV self-testing have improved prevention options, uptake and adherence remain limited [6,7]. This challenge highlights a need for more youth-centred, empathetic, and accessible interventions that can meet young women where they are, socially and digitally.

In recent years, digital health innovations have provided new pathways for HIV prevention and education [8]. With the rapid expansion of mobile connectivity in South Africa, artificial intelligence (AI)-driven chatbots have emerged as promising tools to deliver health information, guide HIV self-testing, and facilitate linkage to services [9–11]. These chatbots offer privacy, convenience, and interactivity qualities that are particularly valuable for young people navigating sensitive topics such as sexual and reproductive health [12,13]. Although digital health interventions for HIV prevention are expanding globally, most studies conducted in South Africa and other resource-limited settings have focused on relatively simple technologies such as short message services (SMS) reminders and basic mobile applications [13,14]. Evidence on more advanced AI-enabled conversational platforms remains limited, with much of the existing literature emphasising feasibility and acceptability rather than behavioural outcomes or differential responsiveness across subgroups [13,14]. Moreover, few studies have examined how behavioural characteristics, such as risk-taking propensity, may influence engagement with AI-driven prevention tools. This gap highlights the need for context-specific evaluations that assess not only overall effectiveness but also variation in responsiveness among diverse groups of AGYW. Limited evidence exists on how these tools influence behavioural intention or how individual differences, such as risk-taking propensity, shape users’ responses to digital interventions.

Risk-taking propensity is a behavioural characteristic that reflects an individual’s tendency to engage in actions with uncertain outcomes despite potential negative consequences [15]. Risk-oriented individuals are more likely to pursue options that offer high potential rewards alongside elevated risks, rather than selecting safer alternatives [15,16]. Their decisions reflect the combined influence of decision characteristics, situational context and individual differences [16]. Among adolescents and young adults, elevated risk-taking has been linked to health-related behaviours such as unprotected sex, substance use and delayed engagement with preventative health services [17–19]. For AGYW in high-HIV burden settings, greater risk-taking may contribute not only to increased exposure to HIV but also to lower perceived vulnerability and reduced motivation to seek prevention or care [18,20,21]. Despite its relevance, risk-taking propensity is rarely measured directly in HIV prevention research, and even less is known about how it influences responsiveness to AI-powered HIV prevention tools.

Risk-taking propensity is a well-established behavioural factor associated with HIV vulnerability among adolescents and young adults [22]. Yet, it is rarely incorporated into evaluations of digital HIV interventions, particularly in low- and middle-income countries (LMICs), where limited resources may hinder a comprehensive evaluation of individual differences [23]. Behavioural tasks such as the Balloon Analogue Risk Task (BART) offer an objective approach to assessing risk-taking propensity that complements self-reported measures and may be especially valuable in settings where social desirability bias and limited health literacy can affect survey responses [24]. The BART is a computer-based behavioural task in which individuals repeatedly decide whether to take incremental risks in exchange for potential rewards, with greater risk-taking reflected in a higher number of risk decisions before a loss occurs [24,25]. By simulating real-world risk reward trade-offs, the BART has been widely used to capture individual differences in risk-taking behaviour across diverse populations [24,25]. Understanding how behavioural characteristics shape responses to digital interventions is critical in resource-limited contexts [24], where scalable tools must effectively engage diverse user profiles to maximise public health impact.

Engagement with HIV services remains a challenge among AGYW, particularly in settings where structural, social and behavioural barriers continue to limit service uptake [26]. Intention to engage in HIV services is an important indicator of readiness and willingness to access prevention and care and is associated with subsequent health seeking behaviour [27]. However, limited evidence exists on factors influencing intention to engage in HIV services following exposure to AI-enabled interventions among AGYW in South Africa. Addressing this gap is important for informing the development of more responsive and effective digital HIV prevention strategies.

Against this backdrop, our study examined whether interaction with an AI-powered conversational tool influenced intention to link to HIV services among AGYW in Gauteng, South Africa. Given evidence that behavioural characteristics shape engagement with health interventions, the study further assessed whether baseline risk-taking propensity moderated changes in intention following exposure to the AI-powered tool. By integrating an objective behavioural measure of risk-taking with a pre-post design, this study aims to contribute evidence on how digital HIV interventions may differentially influence intention to access HIV services among diverse behavioural profiles in a resource-limited setting.

## Methods

### Study Design and Context

In this study, we used data collected as part of the parent study, Self-Care from Anywhere (SCFA). SCFA is a platform that powers AI companions like Aimee via WhatsApp chatbots, leveraging multimodal AI. It provides digital services to support HIV prevention, HIV self-testing guidance, health education, and clinical linkage via WhatsApp. The parent study assessed the usability, acceptability and feasibility of SCFA among adolescent girls and young women in Gauteng Province, South Africa (24). Data collection took place between 18 November 2024 and 30 April 2025.

For this analysis, we employed an observational pre-post design to assess immediate changes in participants’ intention to link to HIV services after engaging with the SCFA platform. The baseline data were collected before SCFA interaction, and follow-up data were collected the same day after interaction with SCFA. BART was administered at baseline, after the pre-intervention survey and before exposure to SCFA. This design enabled measurement of short-term, within-participant changes in intention to link to HIV services following exposure to the AI-powered tool.

### Study Setting

The study was conducted in Gauteng Province, within the Tshwane and Ekurhuleni metropolitan areas. Gauteng is the most densely populated province in South Africa, with substantial socio-economic challenges, including high rates of migration, unemployment estimated at around 39%, and an HIV prevalence of approximately 12% [30,31]. These factors make it an important setting for evaluating innovative HIV prevention strategies targeted at young people.

### Study Population and Recruitment

The target population consisted of AGYW aged 16 to 24 years residing in Gauteng Province. This group represents a key demographic in South Africa’s HIV response, as AGYW experience disproportionately high rates of HIV infection compared to their male counterparts. The primary study aimed to recruit approximately 100 AGYW aged 16 to 24 years, comprising ninety young women aged 18 to 24 years and ten young women aged 16 to 17 years. The larger target for those aged 18 to 24 years reflects the age distribution of individuals routinely reached through service delivery and aligns with the typical age of HIV acquisition in this population. However, given the high HIV vulnerability among adolescent girls and the opportunity to engage them before the usual age of HIV acquisition in South Africa, 16- to 17-year-olds were also included. Recruitment was conducted through Shout-It-Now), that supports youth-focused health and development programs in the province. Participants were recruited using purposive sampling, and potential participants were screened for eligibility during recruitment. Individuals were considered eligible if they self-identified as female, were aged 16-24 years, were able to understand English, and were willing to self-report their HIV status at enrolment. Screening was conducted by Shout-It-Now staff, who completed a screening log documenting eligibility. After recruitment, interested potential participants were transported to the HE^2^RO office in Parktown, Johannesburg, where all study activities took place.

### Measures

The primary independent variable was the intervention period (pre-versus post-intervention), representing exposure to the SCFA tool. The outcome variable was participants’ intention to engage in HIV prevention or treatment services, measured at both time points. Intention for Action (IFA) was measured at baseline and post-intervention using six self-reported items that assessed participants’ intentions to seek HIV-related services and information. IFA reflects early engagement with HIV services and represents a step toward linkage to care. Responses to the six items were summed to create a composite IFA score at each time point, scores ranging from 6 to 40; with higher scores reflecting greater intention and confidence to take HIV-related action. Changes in this score across time points were examined to evaluate the impact of the intervention on participants’ likelihood of accessing HIV services.

Risk-taking propensity was assessed using the BART, a validated behavioural measure of real-world risk-taking. Participants completed 10 rounds in which they inflated a virtual balloon to earn points, balancing potential reward against the risk of the balloon bursting and the loss of accrued points for that round. Risk-taking propensity was operationalised as the mean adjusted number of pumps, calculated by averaging pump counts across rounds in which the balloon did not burst; higher scores indicate greater risk-taking propensity. The BART was administered after completion of the pre-intervention survey and before interaction with the SCFA. Several covariates were included in the analysis to account for potential confounding influences. These covariates included socio-demographic characteristics (participants’ age and education level), behavioural variables (sexual activity status, PrEP use) and HIV status.

### Data Collection

Data collection began with the capture of socio-demographic data using an electronic enrolment form hosted on the REDCap platform. Participants then completed a baseline pre-intervention survey, followed by the BART administered in the Behavioural Laboratory (B-Lab) to objectively measure risk-taking propensity. Following these assessments, participants interacted with the AI-powered tool through WhatsApp. The SCFA answered any health or lifestyle questions, provided confidential guidance on HIV self-testing, empathetic pre- and post-test counselling, clear guidance on next steps and a private channel for engagement and connection with clinicians. Based on their self-reported HIV status, participants living with in HIV only interacted with SCFA whereas a sub-set of participants (n = 41) with a HIV negative diagnosis interacted and engaged in a simulated HIV self-testing process guided by the SCFA. Immediately after the interaction, participants completed a post-intervention survey that repeated the intention-to-engage measures.

### Data Analysis

De-identified pre- and post-intervention data, together with BART scores, were extracted for this secondary analysis. Data were cleaned and analysed using Stata (version 18). Descriptive statistics were calculated for socio-demographic characteristics and BART scores. Histograms were used to visually examine the distributions. To investigate changes in intention before and after the intervention, paired-samples t-tests were conducted. To assess whether risk-taking behaviour moderated changes in intention to engage in HIV services, linear regression models were fitted, including an interaction term between intervention period and BART score.

Multivariable linear regression models were estimated to examine predictors of change in intention to engage in HIV services. All tests were two-sided, with statistical significance reported at alpha levels of 0.05. Variables were entered in successive blocks based on conceptual relevance: socio-demographic characteristics, behavioural variables, health status and an interaction term between the post-intervention indicator and baseline risk-taking propensity to assess moderation. Regression coefficients (β), 95% confidence intervals, and associated *p*-values are reported. Model fit was assessed using the coefficient of determination (R²). Robust standard errors clustered by participants were used to account for repeated observations. External validation was not conducted due to the single-sample design; however, consistency across the multivariate models supports the robustness of findings.

### Ethical Considerations

Ethical approval for the primary study (240906) and for the secondary data analysis (M-250948) was obtained from The University of Witwatersrand Human Research Ethics Committee. All study activities were conducted in accordance with ethical procedures and an approved protocol. Participants aged 18 years and older provided written informed consent on the day of data collection prior to participation. For participants younger than 18 years (minors), written parental consent was obtained at the Shout-It-Now offices prior to inviting potential participants for study participation in the HE^2^RO offices. On the day of data collection, the minors provided written assent to participate in the study. Each participant was assigned a unique study identification number to maintain confidentiality. All data were de-identified before analysis, and participant identifiers were replaced with a unique study ID to preserve confidentiality. Data was stored securely on password-protected servers accessible only to the authorised research team.

## Results

The study sought to enrol 100 AGYW. However, last-minute cancellations by three participants resulted in a smaller final sample. A total of 97 AGYW aged 16 to 24 years participated in the study, including 90 (18–24-year-old) and seven (16-17 years old). Table 1 presents the socio-demographic and baseline characteristics of the entire sample. The median age was 21 years (Interquartile Range [IQR] 19-22), indicating that most participants were older adolescents and young adults. The majority (61.9%) completed high school education (Grade 12). Most participants (63.9%) reported having had sex in the past 12 months and 85.6% self-reported having a HIV-negative status. Nearly a quarter (24.7%) reported current PrEP use, and around one in ten participants preferred not to disclose PrEP use.

**Table 1:**
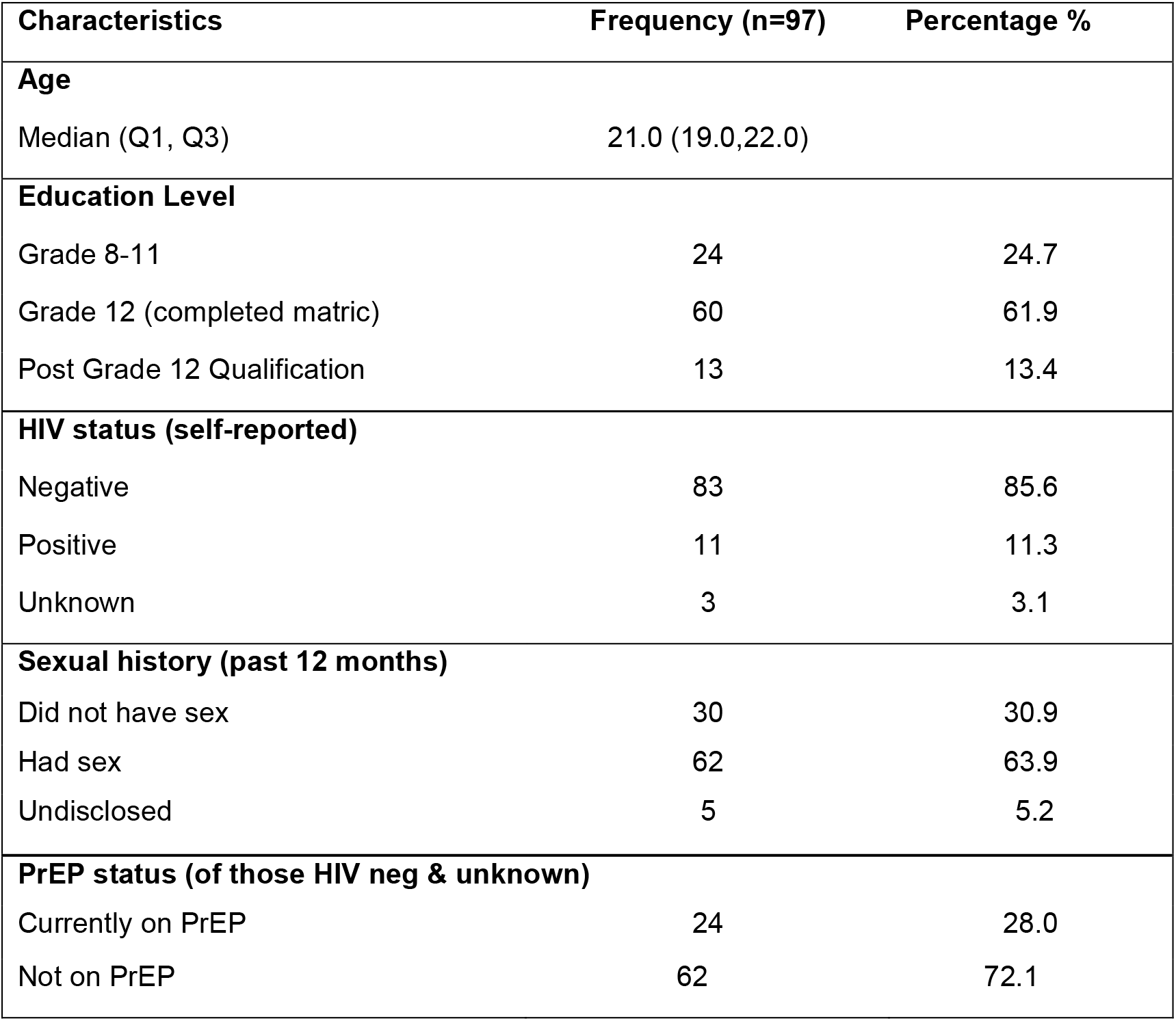
Socio-demographic characteristics of adolescent girls and young women enrolled in Gauteng (N = 97)

| Characteristics | Frequency (n=97) | Percentage % |
|---|---|---|
| **Age** |  |  |
| Median (Q1, Q3) | 21.0 (19.0,22.0) |  |
| **Education Level** |  |  |
| Grade 8-11 | 24 | 24.7 |
| Grade 12 (completed matric) | 60 | 61.9 |
| Post Grade 12 Qualification | 13 | 13.4 |
| **HIV status (self-reported)** |  |  |
| Negative | 83 | 85.6 |
| Positive | 11 | 11.3 |
| Unknown | 3 | 3.1 |
| **Sexual history (past 12 months)** |  |  |
| Did not have sex | 30 | 30.9 |
| Had sex | 62 | 63.9 |
| Undisclosed | 5 | 5.2 |
| **PrEP status (of those HIV neg & unknown)** |  |  |
| Currently on PrEP | 24 | 28.0 |
| Not on PrEP | 62 | 72.1 |

Risk-taking propensity was assessed using the BART and operationalised as the mean adjusted number of pumps per round, with adjusted pump counts calculated only for rounds in which the balloon did not burst (see Table 2). Higher average pump scores indicate greater risk-taking propensity. Participants completed 10 rounds, yielding a mean adjusted pump score of 3.6 pumps per round (SD = 2.5). On average, participants experienced 3.6 balloon bursts per participant (SD = 2.9). Descriptively, the mean adjusted pump count per participant was 18.1 (SD = 9.3).

**Table 2:** Summary statistics of BART performance at baseline among AGYW (N = 97)

| Variable |  | Mean | SD | Minimum | Maximum |
|---|---|---|---|---|---|
| Number of balloons burst | per participant | 3.6 | 2.9 | 0.0 | 10.0 |
| Total pumps count | per participant | 33.1 | 15.3 | 5.0 | 63.0 |
| Total pumps | per round | 3.3 | 1.5 | 0.5 | 6.3 |
| Adjusted pumps count | per participant | 18.1 | 9.3 | 0.0 | 42.0 |
| Adjusted pumps | per round | 3.6 | 2.5 | 1.0 | 10.0 |

The distribution of adjusted pump scores was slightly right skewed, indicating that most participants clustered at the lower end of the observed range, with fewer participants showing higher scores. Figure 1 illustrates the distribution of adjusted pumps, highlighting the spread and shape of baseline risk propensity within the sample. Given the observed variability in baseline risk propensity, we further examined whether intention to engage in HIV services changed following the intervention. A paired-samples t-test was conducted to compare pre- and post-intervention survey IFA scores. The mean pre-intervention IFA score was 22.08 (SD = 2.84), increasing to 22.62 (SD = 2.99) post-intervention. The average increase was 0.54 points (95% CI: −0.11 to 1.19), representing a small increase in intentions. However, there is currently no established minimally important difference for the IFA score, making it difficult to determine the practical significance of this change. Furthermore, the paired t-test indicated that the increase was not statistically significant (t (96) = 1.64, p = 0.105).

**Figure 1.**
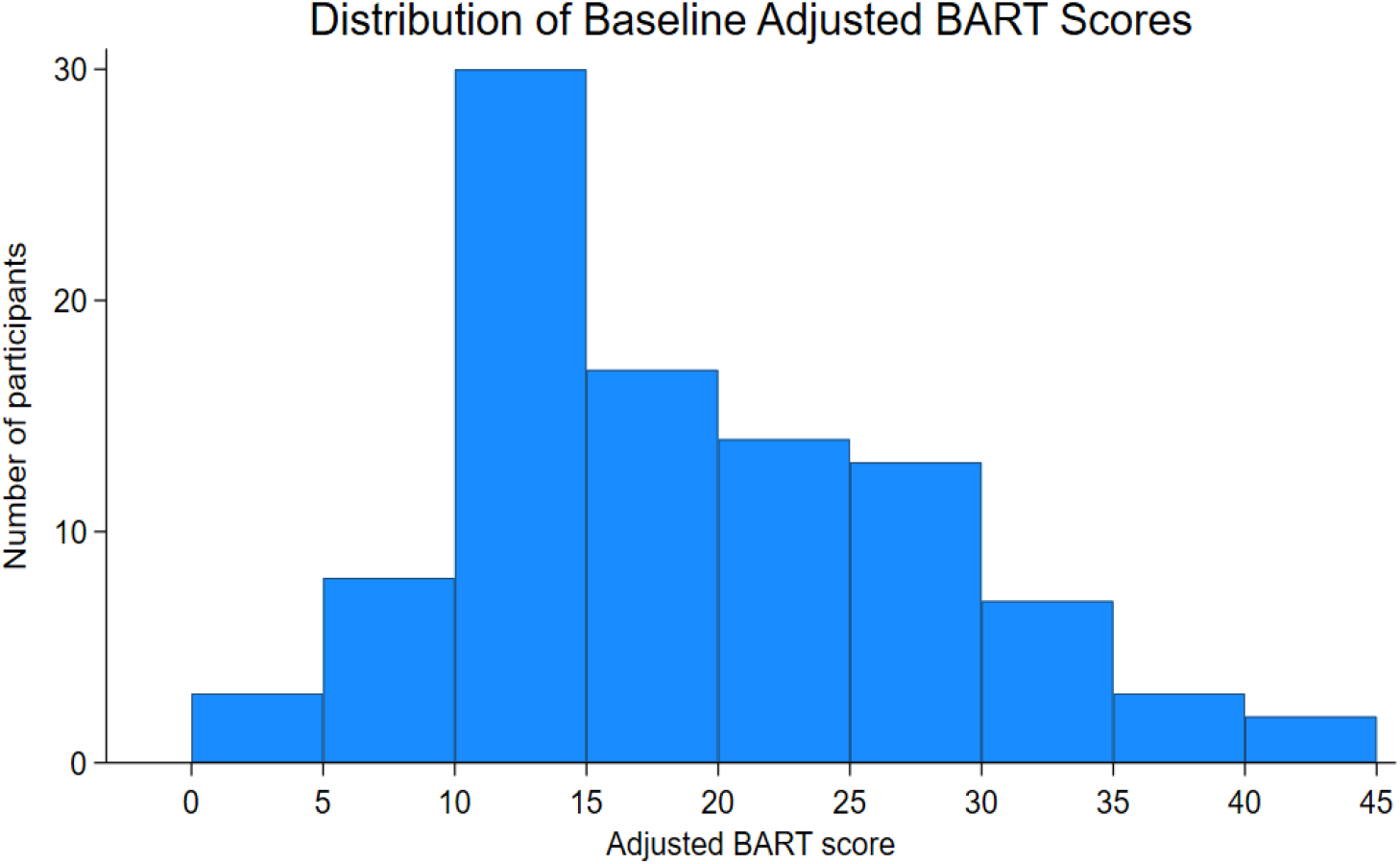
Distribution of baseline adjusted BART pump scores illustrating baseline risk-taking propensity among participants

**Figure 2** presents a box plot of intention to engage in HIV services at the pre- and post-intervention assessments. The figure illustrates the distribution of scores at each time point, including the median, interquartile range, and overall spread. The median post-intervention score appears slightly higher than the pre-intervention score, and the interquartile ranges overlap considerably, indicating that while overall intention increased marginally, variability remained similar across timepoints.

**Figure 2.**
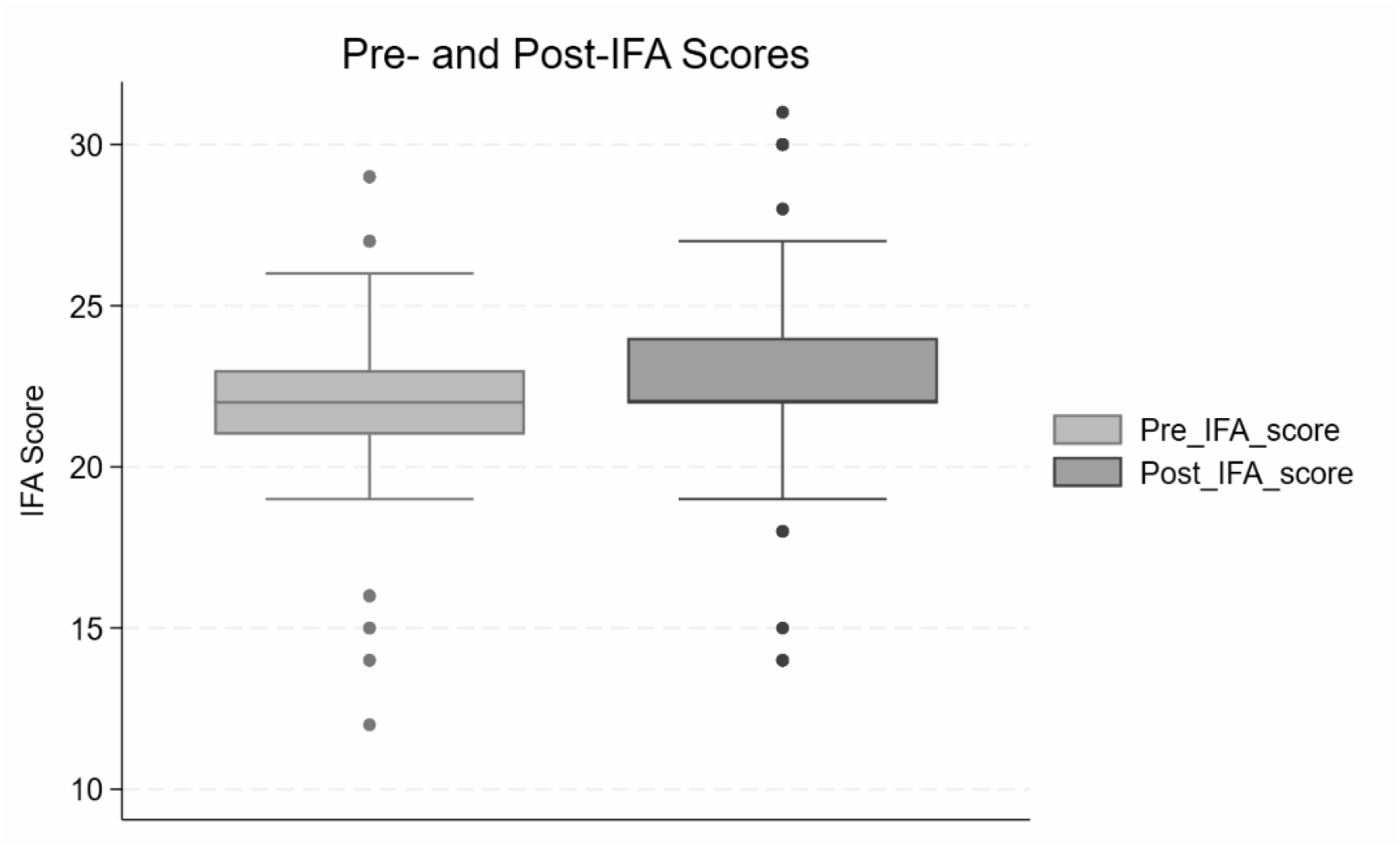
Box plots of pre- and post-intervention IFA scores

To examine whether baseline risk propensity moderated the intervention’s effect on the intention to engage in HIV services, multivariate linear regression models (E1-E7) were estimated, including an interaction term between the post-intervention indicator and baseline risk-taking propensity. Full results for the models are presented in the Supplementary Table S1. Socio-demographic, behavioural and interaction terms were added in successive blocks. We report only on the final fully adjusted model (E7), which directly tested the hypothesised moderation effect while accounting for all covariates. See Table 3. The final model was statistically significant (F (10, 93) = 3.11, p = 0.0019) and accounted for about 17% of the variance in change in intention to engage in HIV services (R² = 0.17). After adjusting for other variables in the model, participants demonstrated an increase in intention score during the post-intervention period compared with pre-intervention period (β = 1.627 (0.625-2.602), p<0.001). A significant interaction was observed between the post-intervention period and baseline risk-taking propensity (β = −0.313, 95% CI: −0.582 to −0.044, p < 0.023), indicating that changes in intentions to engage with HIV services differed according to baseline risk-taking and was lower among risk seeking participants. Each additional year of age was associated with increase in intention to engage with HIV services following interaction with the AI-powered intervention (β = 0.372 (0.050-0.694), p<0.024). AGYW with a self-reported HIV positive status showed significantly lower intention change compared with those who self-reported HIV negative status (β=−2.794 (CI −5.395 – −0.193), p<0.036). Other factors, including education level and recent sexual activity, were not significantly (p>0.05) associated with the change in intention to engage in HIV services.

**Table 3:**
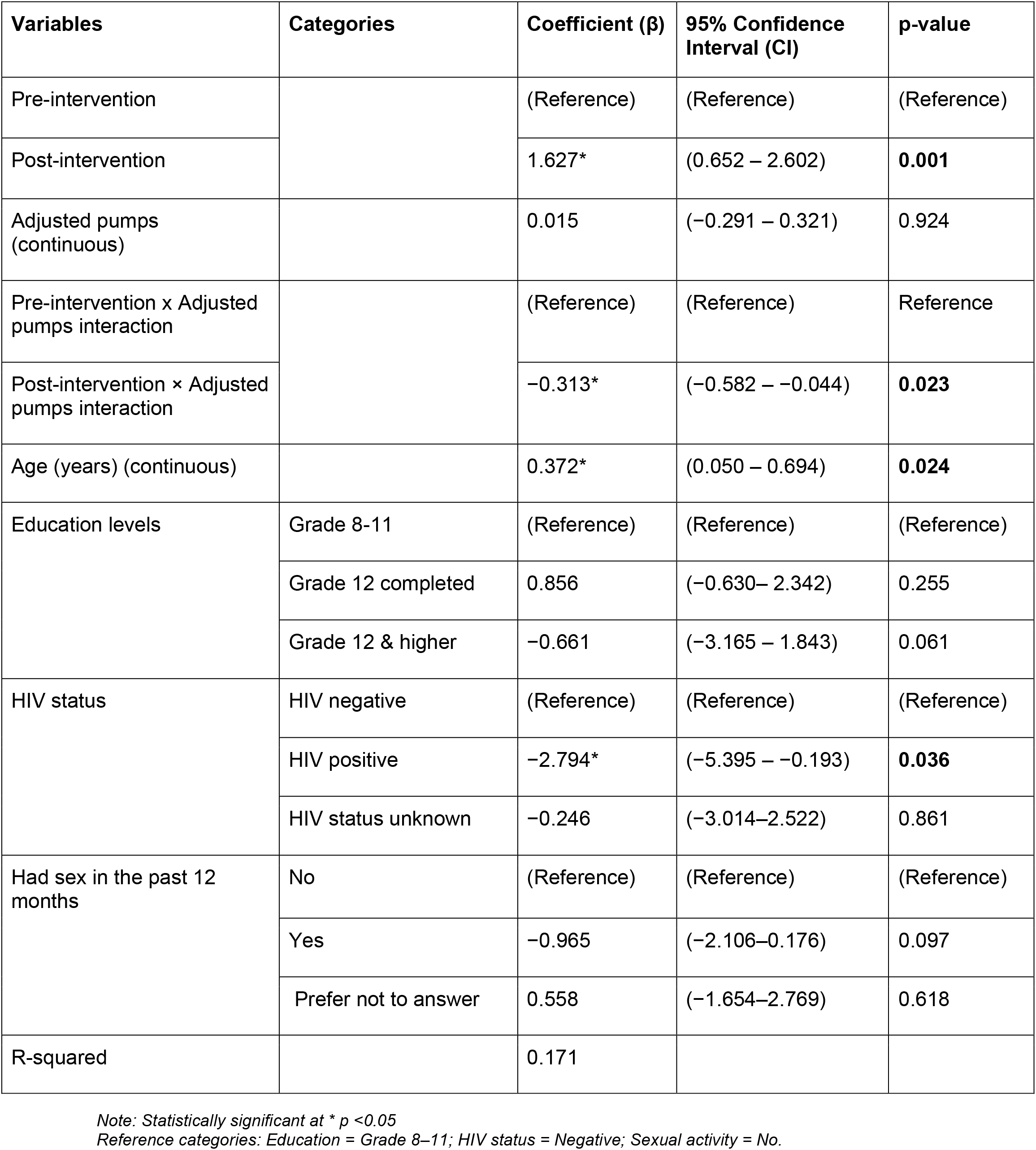
Multivariable linear regression model testing whether baseline risk-taking behaviour moderates the effect of the intervention on the change in intention to engage in HIV services among AGYW aged 16-24 years in Gauteng (N=97)

| **Variables** | **Categories** | **Coefficient (β)** | **95% Confidence Interval (CI)** | **p-value** |
|---|---|---|---|---|
| Pre-intervention |  | (Reference) | (Reference) | (Reference) |
| Post-intervention |  | 1.627* | (0.652 – 2.602) | **0.001** |
| Adjusted pumps (continuous) |  | 0.015 | (−0.291 – 0.321) | 0.924 |
| Pre-intervention x Adjusted pumps interaction |  | (Reference) | (Reference) | Reference |
| Post-intervention × Adjusted pumps interaction |  | −0.313* | (−0.582 – −0.044) | **0.023** |
| Age (years) (continuous) |  | 0.372* | (0.050 – 0.694) | **0.024** |
| Education levels | Grade 8-11 | (Reference) | (Reference) | (Reference) |
|  | Grade 12 completed | 0.856 | (−0.630– 2.342) | 0.255 |
|  | Grade 12 & higher | −0.661 | (−3.165 – 1.843) | 0.061 |
| HIV status | HIV negative | (Reference) | (Reference) | (Reference) |
|  | HIV positive | −2.794* | (−5.395 – −0.193) | **0.036** |
|  | HIV status unknown | −0.246 | (−3.014–2.522) | 0.861 |
| Had sex in the past 12 months | No | (Reference) | (Reference) | (Reference) |
|  | Yes | −0.965 | (−2.106–0.176) | 0.097 |
|  | Prefer not to answer | 0.558 | (−1.654–2.769) | 0.618 |
| R-squared |  | 0.171 |  |  |

*Note: Statistically significant at \* p <0.05*
*Reference categories: Education = Grade 8–11; HIV status = Negative; Sexual activity = No.*

Figure 3 illustrates the interaction between the post-intervention indicator and baseline risk-taking propensity in relation to predicted intention to access HIV services. Predicted intention to engage following the intervention was higher in individuals with lower levels of risk-taking and decreased as risk-taking propensity increased. In contrast, predicted intention to engage before the intervention remained relatively stable across the observed range of risk-taking. This crossover pattern is consistent with the significant interaction term observed in the regression model, indicating that changes in intention following the intervention varied by baseline risk-taking propensity.

**Figure 3.**
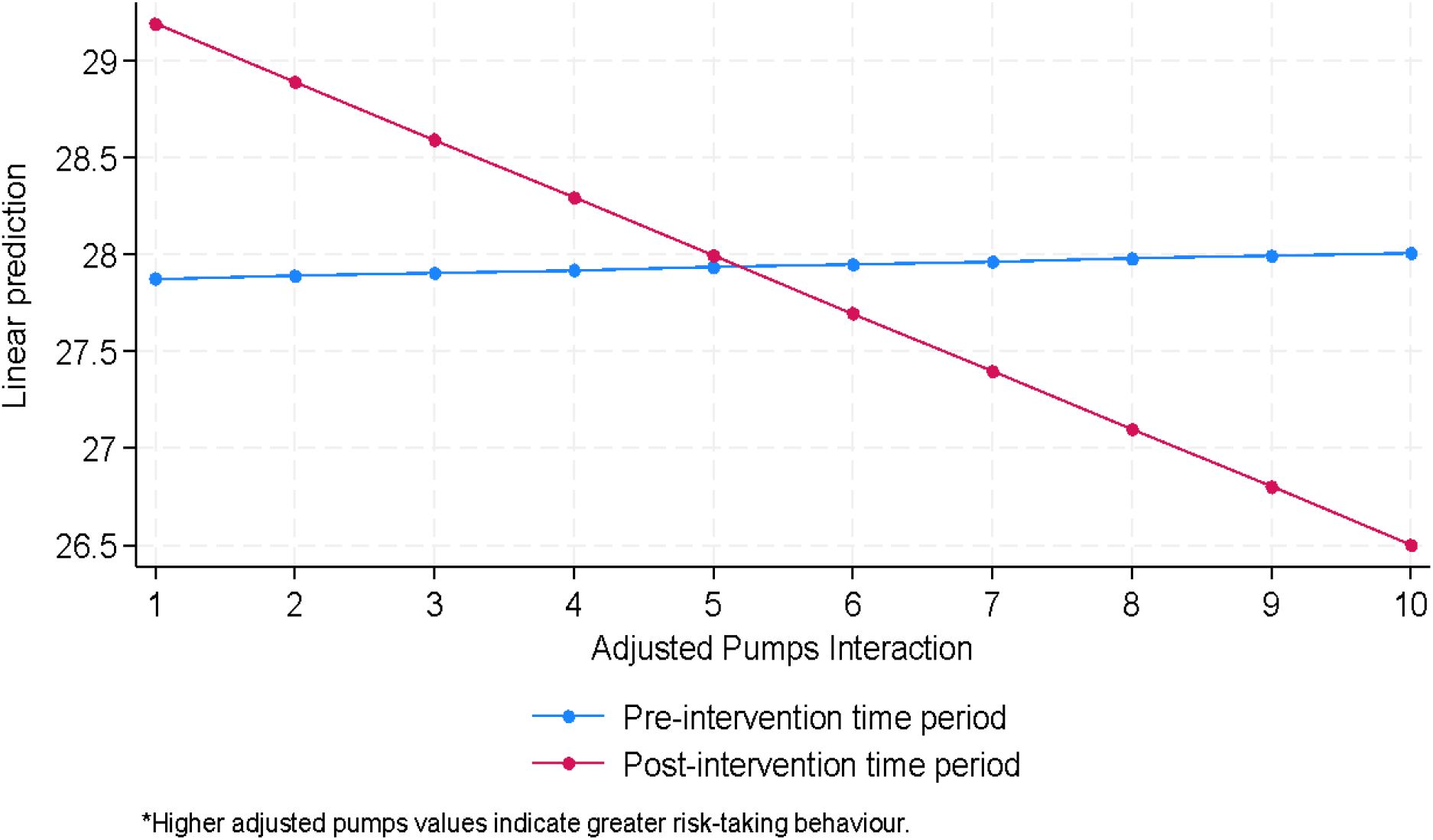
Predicted intention to engage in HIV services across levels of baseline risk-taking propensity before and after the intervention

Overall, the change in intention to engage with HIV services following the intervention was not statistically significant. However, multivariable linear regression and graphical analyses indicated that intervention influence varied by baseline risk-taking propensity. Intentions to engage with HIV services increased among participants who were more risk-averse, whereas this increase was not observed among those with higher baseline risk-taking propensity. These findings suggest that risk-taking propensity maybe an important consideration when examining changes in intentions to engage with HIV services.

## Discussion

Understanding how AI-enabled interventions can support HIV prevention among AGYW in resource-limited settings is increasingly important. This study investigated the relationship between baseline risk-taking propensity and changes in intentions to engage with HIV services among AGYW following interaction with SCFA. This study showed that changes in intentions to engage with HIV services following interaction with an AI-powered tool differed according to baseline risk-taking behaviour among AGYW. Specifically, risk-averse participants demonstrated greater change in intentions to engage with HIV services, compared with risk-seeking participants. These differences became apparent after adjustment for relevant covariates in the multivariable regression model. The variation in baseline BART scores suggests that participants differed in their risk-taking propensity, providing a basis for examining whether these differences were associated with changes in intentions to engage with HIV services. Risk-averse AGYW and older participants demonstrated greater intention to access HIV services than their risk-seeking peers and younger adolescents, respectively.

Although AI-powered interventions have shown promise for supporting HIV prevention, evidence from resource-limited settings remains limited. In South Africa, a resource-limited setting, most existing research has focused on conventional digital interventions such as SMS reminders and basic mobile applications, while more advanced AI-enabled platforms are still largely being evaluated for feasibility and acceptability [14,33,34]. This is particularly important given that AGYW continue to experience a disproportionately high burden of HIV infection, yet uptake of effective prevention strategies such as PrEP remains low [6,7]. Improving engagement with HIV services, including PrEP, is therefore a public health priority. The findings of this study suggest that AI-powered interventions may represent a promising approach to supporting engagement with HIV prevention services but also highlight that responses may differ according to baseline risk-taking behaviour. Future research should therefore investigate how AI-powered interventions can be tailored to the needs of both risk-averse and risk-seeking AGYW to improve engagement across the HIV prevention cascade.

Risk-taking behaviour may influence how AGYWs engage with digital HIV interventions. The observed differences suggest that baseline risk-taking behaviour is an important behavioural characteristic to consider when designing AI-powered HIV interventions, as AGYW with different risk profiles may respond differently to the same intervention. Literature shows that risk-averse individuals tend to be more responsive to potential negative outcomes and are therefore more likely to adopt protective behaviours when presented with health information [35,36]. In contrast, those who are more risk-seeking often place greater value on immediate rewards and may downplay future risks, which can limit the extent to which their intentions shift [16]. Given that AGYW with higher baseline risk-taking behaviour showed smaller improvements in intentions to engage in HIV services, future AI-powered interventions may need to incorporate tailored strategies that better address the needs and decision-making patterns of this group, while maintaining approaches that are effective for more risk-averse individuals. In resource-limited settings, where digital interventions are increasingly adopted to broaden access to HIV services, considering differences in baseline risk-taking behaviour may be particularly important.

Behavioural risk-taking is a key determinant of health-related decision-making and is particularly relevant in HIV prevention among young women. Behavioural differences in willingness to take risks may influence how health information is interpreted and acted upon, highlighting the importance of considering these differences when designing digital HIV prevention strategies [16]. Existing literature shows that adolescent risk-taking is shaped by developmental, social, and contextual factors, and that higher BART scores are associated with greater real-world risk behaviours [37–39]. By characterising these behavioural risk profiles, this study adds to the evidence by documenting the distribution of risk-taking propensity among AGYW in a resource-limited setting, where such data remain limited. In the context of AI-enabled interventions, such variation in risk-taking propensity highlights the need to consider behavioural diversity when designing strategies to support engagement with HIV services.

Beyond risk-taking propensity, age-related differences highlight how developmental factors shape engagement with digital HIV interventions. Participants toward the upper end of the age range demonstrated greater intention to access HIV services, possibly reflecting increased autonomy, stronger self-efficacy, and greater confidence in navigating health systems as young women transition into adulthood [40]. This finding aligns with prior research showing that older AGYW are more likely to independently access sexual and reproductive health services and report higher levels of perceived behavioural control in health decision-making [41]. This extends existing evidence by showing how these age-related differences shape responsiveness to HIV interventions. However, younger adolescents often face structural and relational constraints, including limited decision-making power, dependence on caregivers, and concerns about confidentiality, which may limit their responsiveness to digital prompts [41,42]. These challenges may be further amplified in resource-limited settings, where access to youth-friendly and confidential services may already be constrained. These findings suggest that age-sensitive digital design and tailored engagement strategies are important for supporting equitable engagement across the adolescent age spectrum, particularly as digital HIV interventions are scaled and implemented in resource-limited settings.

These insights have important implications for how AI-enabled HIV interventions are designed and implemented. Taken together, these findings indicate that AI-enabled interventions hold considerable promise but are unlikely to achieve equitable impact without behaviourally tailored design. Strengthening responsiveness among AGYW with higher risk-taking propensity may require personalised content, adaptive message framing, and engagement features that explicitly address risk perception and motivational drivers. Previous studies found that as digital health platforms continue to expand within HIV prevention and care, integrating behavioural science principles into AI system architecture will be critical to ensuring that interventions do not disproportionately benefit those already oriented toward service engagement [43,44]. In resource-limited LMIC settings, where in-person counselling capacity and differentiated service delivery models are often constrained, strategically designed AI-enabled tools could offer a scalable and potentially cost-efficient mechanism to enhance targeted linkage and optimize prevention cascade outcomes.

These findings highlight the importance of considering risk-taking propensity in future research and the development of AI-powered HIV interventions to ensure they are responsive to the diverse needs of AGYW. In the South African context, where PrEP uptake among AGYW remains below desired levels, incorporating behavioural traits such as risk-taking propensity into intervention design may help inform more targeted strategies to improve engagement with HIV prevention services.

Notwithstanding these contributions, limitations and several strengths should be considered when interpreting the findings. The observational pre-post design without a control group limits causal inference, as changes in intention cannot be definitively attributed to the intervention. Although interaction effects were examined, subgroup analyses may have been underpowered, particularly for participants living with HIV, and should therefore be interpreted with caution. Age was treated as a continuous variable because the small number of participants aged 16-17 years (n = 7) limited meaningful age-group comparisons. Intention to engage in HIV services may not necessarily translate into actual service uptake. The study did not collect information on whether participants living with HIV were engaged in HIV care or receiving treatment. Consequently, it was not possible to determine their level of engagement in care or establish how this may have influenced their intention for action. In addition, the relatively short follow-up period precluded assessment of sustained behavioural change over time. Finally, interaction with the SCFA took place within a structured study environment, which may differ from real-world, unsupervised implementation conditions.

However, this study has several strength worth noting. A key strength of this study is the use of the BART as an objective measure of behavioural risk-taking propensity, moving beyond the self-reported indicators commonly used in HIV prevention research. The explicit examination of moderation within a digital HIV intervention context strengthens the analysis by identifying subgroup-specific differences in responsiveness. The use of multivariable regression modelling allowed for the adjustment of relevant covariates while testing interaction effects, providing insight beyond average treatment estimates. Importantly, by examining intention to engage in HIV services rather than solely acceptability or usability outcomes, this study addresses a critical intermediate step in the HIV prevention cascade. This study offers insight into how behavioural and developmental factors may shape changes in intention to engage in HIV services among AGYW following interaction with an AI-enabled intervention.

## Conclusions

Persistent gaps in engagement with HIV services among AGYW highlight the need for innovative and responsive digital approaches. This study suggests that responsiveness to AI-enabled HIV interventions varies by behavioural and developmental factors. AI-powered conversational tools may strengthen the intention to engage in HIV services among AGYW, if designed to accommodate diverse behavioural profiles. Digital interventions aimed at improving engagement should account for variation in readiness, risk orientation, and lived experience to prevent uneven engagement across subgroups. Using behavioural insights to design and evaluate AI-powered platforms can help move beyond one-size-fits-all approaches and become more responsive to the factors shaping AGYW’s health decision-making. As HIV prevention strategies increasingly incorporate digital technologies, ensuring that these tools are behaviourally adaptive and equity-focused will be important for supporting more inclusive engagement with HIV services, particularly for young women in resource-limited settings.

## Supporting Information

**Supplementary Table 1 (S1):**
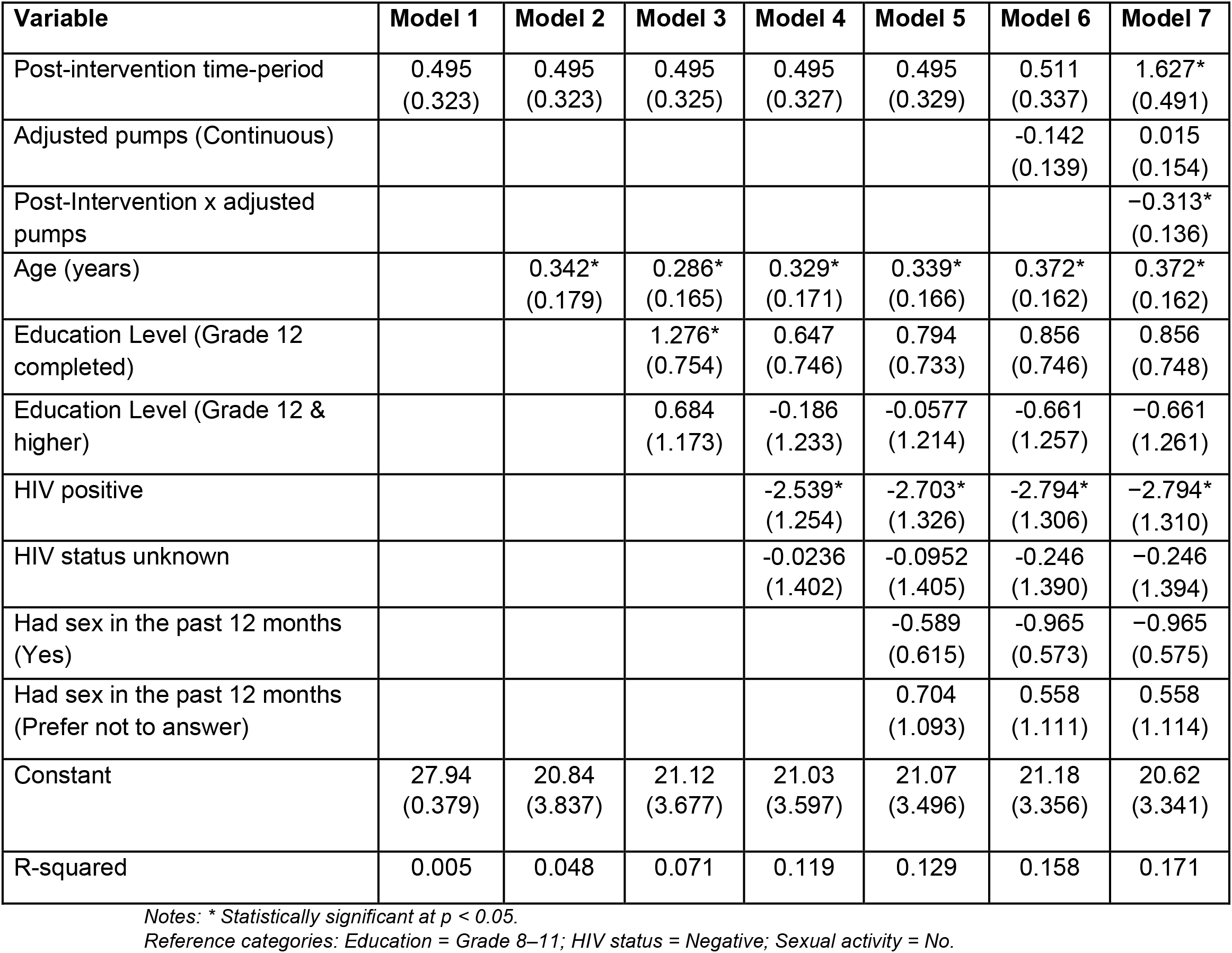
Multivariate linear regression models (E1-E7) predicting change in intention to link to HIV services among AGYW.

## Ethics approval

The study was approved by ethics committees at the University of the Witwatersrand for main-study (ref: 240906) and sub-study (ref: 250948) and Boston University (H-45383).

## Data Availability

Data are available on request to the corresponding author.

## Acknowledgements

We would like to thank Shout-It-Now for their support in recruiting participants and to Ms. Busisiwe Sibiya for leading data collection and Mr Ross Greener for assisting with the Ballon Analogue Risk Task activities. We sincerely thank all participants for generously sharing their time and insights, which made this study possible.

## References

1. UNAIDS. HIV and adolescent girls and young women - Thematic briefing note - 2024 global AIDS update The Urgency of Now: AIDS at a Crossroads [Internet]. 2024 [cited 2025 Dec 4]. Available from: https://aidsinfo.unaids.org/

2. Murewanhema G, Musuka G, Moyo P, Moyo E, Dzinamarira T. HIV and adolescent girls and young women in sub-Saharan Africa: A call for expedited action to reduce new infections. IJID Regions. 2022 Dec 1;5:30. doi:10.1016/J.IJREGI.2022.08.009 PubMed PMID: 36147901.

3. UNAIDS. Global HIV statistics [Internet]. 2025 [cited 2026 Feb 8]. Available from: https://www.unaids.org/sites/default/files/2025-07/2025_Global_HIV_Factsheet_en.pdf

4. Pleaner M, Scorgie F, Martin C, Butler V, Muhwava L, Mojapele M, et al. Introduction and integration of PrEP and sexual and reproductive health services for young people: Health provider perspectives from South Africa. Frontiers in Reproductive Health. 2022 Jan 9;4:1086558. doi:10.3389/frph.2022.1086558

5. Dzinamarira T, Moyo E, Murewanhema G. Navigating the Complexities of HIV Prevention for Adolescents and Young Persons: A Science-to-Program and Systems Approach [Internet]. 2025. doi:10.3390/adolescents5010001

6. Rousseau E, Julies RF, Madubela N, Kassim S. Novel Platforms for Biomedical HIV Prevention Delivery to Key Populations — Community Mobile Clinics, Peer-Supported, Pharmacy-Led PrEP Delivery, and the Use of Telemedicine. Curr HIV/AIDS Rep. 2021 Dec 1;18(6):500. doi:10.1007/S11904-021-00578-7 PubMed PMID: 34708316.

7. Nyblade L, Ndirangu JW, Speizer IS, Browne FA, Bonner CP, Minnis A, et al. Stigma in the health clinic and implications for PrEP access and use by adolescent girls and young women: conflicting perspectives in South Africa. BMC Public Health. 2022 Dec 1;22(1):1916. doi:10.1186/S12889-022-14236-Z PubMed PMID: 36242000.

8. van Heerden A, Bosman S, Swendeman D, Comulada WS. Chatbots for HIV Prevention and Care: a Narrative Review. Curr HIV/AIDS Rep. 2023 Dec 1;20(6):481–6. doi:10.1007/S11904-023-00681-X PubMed PMID: 38010467.

9. Phatsoane Gaven M, Quaife M, Majam M, Singh L, Rhagnath N, Wonderlik T, et al. HIV self-test reporting using mHealth platforms: A pilot study in Johannesburg, South Africa. Frontiers in Reproductive Health. 2023;5. doi:10.3389/FRPH.2023.1073492

10. Hlongwa M, Moyo E, Dzinamarira T. Approaches for improving linkage to HIV care among HIV self-testing individuals in sub-Saharan Africa. BMJ Glob Health. 2023 Jul 14;8(7):e012664. doi:10.1136/BMJGH-2023-012664 PubMed PMID: 37451688.

11. Gous N, Fischer AE, Rhagnath N, Phatsoane M, Majam M, Lalla-Edward ST. Evaluation of a mobile application to support HIV self-testing in Johannesburg, South Africa. South Afr J HIV Med. 2020 Jun 30;21(1). doi:10.4102/SAJHIVMED.V21I1.1088 PubMed PMID: 32670629.

12. Mills R, Mangone ER, Lesh N, Mohan D, Baraitser P. Chatbots to Improve Sexual and Reproductive Health: Realist Synthesis. J Med Internet Res. 2023 Jan 1;25:373–83. doi:10.2196/46761 PubMed PMID: 37556194.

13. Ntinga X, Musiello F, Keter AK, Barnabas R, van Heerden A. The Feasibility and Acceptability of an mHealth Conversational Agent Designed to Support HIV Self-testing in South Africa: Cross-sectional Study. J Med Internet Res. 2022 Dec 1;24(12):e39816. doi:10.2196/39816 PubMed PMID: 36508248.

14. Mahlalela NB, Maboa O, Govathson C, Rossouw L, Long L, Greener R, et al. Evaluating the acceptability, usability and clinical appropriateness of Your Path, an AI-powered tool facilitating relevant access to HIV services post-HIV self-testing in South Africa. 2025. doi:10.1101/2025.11.10.25339576

15. Saleme DM, Moustafa AA. The multifaceted nature of risk-taking in drug addiction. Cognitive, Clinical, and Neural Aspects of Drug Addiction. 2020 Jan 1;41–60. doi:10.1016/B978-0-12-816979-7.00003-0

16. de-Juan-Ripoll C, Chicchi Giglioli IA, Llanes-Jurado J, Marín-Morales J, Alcañiz M. Why Do We Take Risks? Perception of the Situation and Risk Proneness Predict Domain-Specific Risk Taking. Front Psychol. 2021 Mar 8;12:562381. doi:10.3389/fpsyg.2021.562381

17. Promoting Positive Adolescent Health Behaviors and Outcomes. Promoting Positive Adolescent Health Behaviors and Outcomes. 2020 Feb 21. doi:10.17226/25552

18. HIV and sexual risk behaviours by 18-25-year-old youth at Nyandeni Municipality in the Eastern Cape | Mntonintshi-Mketo | Health SA Gesondheid [Internet]. [cited 2025 Apr 8]. Available from: https://hsag.co.za/index.php/hsag/article/view/2541/html

19. Brooks M, Nguyen R, Bruno R, Peacock A. A systematic review of event-level measures of risk-taking behaviors and harms during alcohol intoxication. Addictive Behaviors. 2019 Dec 1;99. doi:10.1016/j.addbeh.2019.106101 PubMed PMID: 31473569.

20. Ninsiima LR, Chiumia IK, Ndejjo R. Factors influencing access to and utilisation of youth-friendly sexual and reproductive health services in sub-Saharan Africa: a systematic review. Reprod Health. 2021 Jun 27;18(1):135. doi:10.1186/S12978-021-01183-Y/TABLES/1 PubMed PMID: 34176511.

21. Kwena ZA, Amico RK, Masvawure TB, Ngure KK, Bukusi EA, Remien RH, et al. Barriers to linkage and retention in HIV care still persist among adolescent girls and young women in western Kenya. African Journal of AIDS Research. 2023;22(2):71–84. doi:10.2989/16085906.2023.2197879 PubMed PMID: 37337818.

22. Majelantle RG, Bainame K, McLaren Z. Knowledge and risky behaviours associated with the spread of HIV/AIDS among young persons in Botswana. Botswana Notes and Records. 2010;42:121–133. JSTOR [Internet]. [cited 2026 Feb 8]. Available from: https://www.jstor.org/stable/23237977

23. Bose DL, Hundal A, Singh S, Singh S, Seth K, Hadi S ul, et al. Evidence and gap map report: Social and Behavior Change Communication (SBCC) interventions for strengthening HIV prevention and research among adolescent girls and young women (AGYW) in low- and middle-income countries (LMICs). Campbell Systematic Reviews. 2023 Mar 1;19(1):e1297. doi:10.1002/cl2.1297

24. Lejuez CW, Read JP, Kahler CW, Richards JB, Ramsey SE, Stuart GL, et al. Evaluation of a behavioral measure of risk taking: The Balloon Analogue Risk Task (BART). Journal of Experimental Psychology: Applied. 2002;8(2):75–84.

25. MacLean RR, Pincus AL, Smyth JM, Geier CF, Wilson SJ. Extending the Balloon Analogue Risk Task to Assess Naturalistic Risk Taking via a Mobile Platform. J Psychopathol Behav Assess. 2017 Mar 1;40(1):107. doi:10.1007/s10862-017-9628-4 PubMed PMID: 30505069.

26. Duby Z, Bunce B, Fowler C, Jonas K, Bergh K, Govindasamy D, et al. These girls have a chance to be the future generation of HIV negative: experiences of implementing a PrEP programme for adolescent girls and young women in South Africa. AIDS Behav. 2023 Jan;27(1):134–149. doi:10.1007/s10461-022-03750-1.

27. Keene CM, Euvrard J, Amico KR, Ragunathan A, English M, McKnight J, et al.; InCARE Stakeholder Group. Conceptualising engagement with HIV care for people on treatment: the Indicators of HIV Care and AntiRetroviral Engagement (InCARE) Framework. BMC Health Serv Res. 2023 May 4;23(1):435. doi:10.1186/s12913-023-09433-4.

28. Shirdel F, Mobasheri N, Kaveh MH, Hassanzadeh J, Ghahremani L. A Randomized Controlled Trial to Evaluate the Effectiveness of a Theory of Planned Behavior-Based Educational Intervention in Reducing Internet Addiction Among Adolescent Girls in Southern Iran. Adolescents 2025, Vol 5, Page 33. 2025 Jul 11;5(3):33. doi:10.3390/adolescents5030033

29. Kakoko DC, Åstrøm AN, Lugoe WL, Lie GT. Predicting intended use of voluntary HIV counselling and testing services among Tanzanian teachers using the theory of planned behaviour. Soc Sci Med. 2006 Aug 1;63(4):991–9. doi: 10.1016/j.socscimed.2006.02.016 PubMed PMID: 16603299.

30. SANAC. Implementation framework to address and monitor social and structural drivers of HIV/AIDS, TB, & STIs amongst Adolescent Girls and Young Women (AGYW). 2022 Sep.

31. HSRC. SABSSM VI Provincial Dialogue: Gauteng Province Media Pack - HSRC [Internet]. 2024 [cited 2025 Nov 11]. Available from: https://hsrc.ac.za/special-projects/sabssm-survey-series/sabssm-vi-provincial-dialogue-gauteng-province-media-pack/

32. Bokolo S, Govathson C, Rossouw L, Madlala S, Frade S, Cooper S, et al. Self-care from anywhere: Evaluating the usability of an AI-powered HIV toolkit among adolescent girls and young women and healthcare providers in South Africa [Preprint]. 2026 Apr 2.

33. Fischer AE, Hanif H, Stocks JB, Rochelle AE, Dominguez K, Armora Langoni EG, et al. Mobile Health Intervention Tools Promoting HIV Pre-Exposure Prophylaxis Among Adolescent Girls and Young Women in Sub-Saharan Africa: Scoping Review. JMIR Mhealth Uhealth. 2025;13:e60819. doi:10.2196/60819 PubMed PMID: 40540732.

34. Shaluhiyah Z, Qatrannada SA, Kusumawati A, Miah MS. The effect of digital health intervention in promoting healthy behavior: A systematic scoping review on strategies to prevent non-communicable diseases. Dialogues in Health. 2025 Dec 1;7:100258. doi:10.1016/j.dialog.2025.100258 PubMed PMID: 41328330.

35. Petrocchi S, Iannello P, Ongaro G, Antonietti A, Pravettoni G. The interplay between risk and protective factors during the initial height of the COVID-19 crisis in Italy: The role of risk aversion and intolerance of ambiguity on distress. Current Psychology. 2022 Jan;41:437–48. doi:10.1007/s12144-021-01601-1

36. Cerami C, Galandra C, Santi GC, Dodich A, Cappa SF, Vecchi T, et al. Risk-Aversion for Negative Health Outcomes May Promote Individual Compliance to Containment Measures in Covid-19 Pandemic. Front Psychol. 2021;12:666454. doi:10.3389/fpsyg.2021.666454 PubMed PMID: 34220639.

37. Meisel SN, Colder CR. Adolescent Social Norms and Alcohol Use: Separating Between- and Within-Person Associations to Test Reciprocal Determinism. Journal of Research on Adolescence. 2020 Feb;30:499–515. doi:10.1111/jora.12494 PubMed PMID: 30908811.

38. Habib M, Osmont A, Tavani JL, Cassotti M, Caparos S. Is adolescence believed to be a period of greater risk taking than adulthood? Int J Adolesc Youth. 2023;28. doi:10.1080/02673843.2023.2242469

39. Duell N, Steinberg L, Icenogle G, Chein J, Chaudhary N, Di Giunta L, et al. Age Patterns in Risk Taking Across the World. J Youth Adolesc. 2018 May;47:1052–72. doi:10.1007/s10964-017-0752-y PubMed PMID: 29047004.

40. Warzywoda S, Fowler JA, Dyda A, Fitzgerald L, Mullens AB, Dean JA. Pre-exposure prophylaxis access, uptake and usage by young people: a systematic review of barriers and facilitators. Ther Adv Infect Dis. 2024;11:20499361241303416. doi:10.1177/20499361241303415 PubMed PMID: 39650691.

41. Muchanga V, Huo L, Kampa KT, Chilundo B, Munguambe K, Moon TD. Knowledge and experiences of adolescent girls and young women in the use of sexual reproductive health and HIV services at health facilities in Maputo City, Mozambique. Frontiers in reproductive health. 2025;7:1667930. doi:10.3389/frph.2025.1667930 PubMed PMID: 41358263.

42. Duby Z, Verwoerd W, Isaksen K, Jonas K, Maruping K, Dietrich J, et al. ‘I can’t go to her when I have a problem’: sexuality communication between South African adolescent girls and young women and their mothers. Sahara J. 2022;19:8–21. doi:10.1080/17290376.2022.2060295 PubMed PMID: 35443869.

43. Mbewe A, Sarkwah HN, Ijaodola O, Ratevosian J, Simba S, Smith TR, et al. The role of digital health and artificial intelligence in improving the reach and effectiveness of HIV prevention in Africa. The Lancet. Global health. 2026. p. e131–42. doi:10.1016/S2214-109X(25)00447-4 PubMed PMID: 41349551.

44. Burrus O, Gupta C, Ortiz A, Zulkiewicz B, Furberg R, Uhrig J, et al. Principles for Developing Innovative HIV Digital Health Interventions. Medical Care. Lippincott Williams and Wilkins; 2018. p. 756–60. doi:10.1097/MLR.0000000000000957 PubMed PMID: 3000125

