## Supplemental Table for "Assessing intention of linking to HIV services among female youth with diverse risk-taking profiles after interacting with an AI-powered tool in Gauteng"

**Supporting Information**

**Supplementary Table 1 (S1):** Multivariate linear regression models (E1-E7) predicting change in intention to link to HIV services among AGYW

| **Variable** | **Model 1** | **Model 2** | **Model 3** | **Model 4** | **Model 5** | **Model 6** | **Model 7** |
|---|---|---|---|---|---|---|---|
| Post-intervention time-period | 0.495 (0.323) | 0.495 (0.323) | 0.495 (0.325) | 0.495 (0.327) | 0.495 (0.329) | 0.511 (0.337) | 1.627* (0.491) |
| Adjusted pumps (Continuous) | | | | | | -0.142 (0.139) | 0.015 (0.154) |
| Post-Intervention x adjusted pumps | | | | | | | −0.313* (0.136) |
| Age (years) | | 0.342* (0.179) | 0.286* (0.165) | 0.329* (0.171) | 0.339* (0.166) | 0.372* (0.162) | 0.372* (0.162) |
| Education Level (Grade 12 completed) | | | 1.276* (0.754) | 0.647 (0.746) | 0.794 (0.733) | 0.856 (0.746) | 0.856 (0.748) |
| Education Level (Grade 12 & higher) | | | 0.684 (1.173) | -0.186 (1.233) | -0.0577 (1.214) | -0.661 (1.257) | −0.661 (1.261) |
| HIV positive | | | | -2.539* (1.254) | -2.703* (1.326) | -2.794* (1.306) | −2.794* (1.310) |
| HIV status unknown | | | | -0.0236 (1.402) | -0.0952 (1.405) | -0.246 (1.390) | −0.246 (1.394) |
| Had sex in the past 12 months (Yes) | | | | | -0.589 (0.615) | -0.965 (0.573) | −0.965 (0.575) |
| Had sex in the past 12 months (Prefer not to answer) | | | | | 0.704 (1.093) | 0.558 (1.111) | 0.558 (1.114) |
| Constant | 27.94 (0.379) | 20.84 (3.837) | 21.12 (3.677) | 21.03 (3.597) | 21.07 (3.496) | 21.18 (3.356) | 20.62 (3.341) |
| R-squared | 0.005 | 0.048 | 0.071 | 0.119 | 0.129 | 0.158 | 0.171 |

*Notes: * Statistically significant at $p < 0.05$.*
*Reference categories: Education = Grade 8–11; HIV status = Negative; Sexual activity = No.*
